# Improved detection of antibody against SARS-CoV-2 by microsphere-based antibody assay

**DOI:** 10.1101/2020.05.26.20113191

**Authors:** Carol Ho-Yan Fong, Jian-Piao Cai, Thrimendra Kaushika Dissanayake, Lin-Lei Chen, Charlotte Yee-Ki Choi, Lok-Hin Wong, Anthony Chin-Ki Ng, Polly K.P. Pang, Deborah Tip-Yin Ho, Rosana Wing-Shan Poon, Tom Wai-Hin Chung, Siddharth Sridhar, Kwok-Hung Chan, Jasper Fuk-Woo Chan, Ivan Fan-Ngai Hung, Kwok-Yung Yuen, Kelvin Kai-Wang To

## Abstract

**Objective:** Currently available COVID-19 antibody tests using enzyme immunoassay (EIA) or immunochromatographic assay have variable sensitivity and specificity. Here, we developed and evaluated a novel microsphere-based antibody assay (MBA) for the detection of immunoglobulin G (IgG) against SARS-CoV-2 nucleoprotein (NP) and spike protein receptor binding domain (RBD).

**Method:** We developed a microsphere-based assay (MBA) to determine the levels of IgG against SARS-CoV-2 NP and spike RBD. The seropositive cut-off mean fluorescent intensity (MFI) was set using a cohort of 294 anonymous serum specimens collected in 2018. The specificity was assessed using serum specimens collected from organ donors or influenza patients before 2020. Seropositive rate was determined among patients with COVID-19. Time-to-seropositivity and signal-to-cutoff (S/CO) ratio were compared between MBA and EIA.

**Results:** MBA had a specificity of 100% (93/93; 95% confidence interval [CI], 96-100%) for anti-NP IgG and 98.9% (92/93; 95% CI 94.2-100%) for anti-RBD IgG. The MBA seropositive rate for convalescent serum specimens of COVID-19 patients were 89.8% (35/39) for anti-NP IgG and 79.5% (31/39) for anti-RBD IgG. The time-to-seropositivity was shorter with MBA than that of EIA. When compared with EIA, MBA could better differentiate between COVID-19 patients and negative controls with significantly higher S/CO ratio for COVID-19 patients and lower S/CO ratio with negative controls. MBA also had fewer specimens in the equivocal range (S/CO 0.9-1.1) than EIA.

**Conclusion:** MBA is robust and simple, and is suitable for clinical microbiology laboratory for the accurate determination of anti-SARS-CoV-2 antibody for retrospective diagnosis, serosurveillance, and vaccine trials.

## INTRODUCTION

In 2003, severe acute respiratory syndrome coronavirus (SARS-CoV) has caused the first severe coronavirus epidemic, leading to more than 8000 cases mainly in Asia *^1,2^*. In 2019, a novel coronavirus, now known as the severe acute respiratory syndrome coronavirus 2 (SARS-CoV-2) has become the first coronavirus to cause a global pandemic ^3^. Unlike the 2003 SARS-CoV, the novel SARS-CoV-2 transmits efficiently among humans, possibly due to high viral load at presentation ^4^ and efficient binding to the human receptor angiotensin receptor 2.

Antibody assays play a major role in clinical management, contact tracing, vaccine studies and the understanding of the epidemiology and pathogenesis of COVID-19 ^5,6^ Antibody testing allows the retrospective diagnosis of an infection by comparing the antibody titer at the acute and at the convalescent phase of the illness. This is especially important for patients whose viral load is too low to be detected by virus detection assays. Furthermore, antibody testing is the preferred method for identifying subclinical infections.

Several assays have been developed to detect antibody against SARS-CoV-2. Enzyme immunoassay is a commonly used antibody assay for the detection of SARS-CoV-2 ‘. We have previously used enzyme immunoassay to determine the serial antibody profile of COVID-19 patients ^7^ and to determine the seroprevalence of SARS-CoV-2 in Hong Kong and in Hubei province ^10^. Lateral flow immunochromatographic assay allows rapid detection, but Currently available antibody testing assays for SARS-CoV-2 mainly relies on enzyme immunoassay or lateral flow immunochromatographic assaysm but the sensitivities of these assays are relatively low ^11^.

With the advance in technology, microsphere-based antibody assay (MBA) using flow cytometers have been developed for different clinical applications. The use of multiplex microsphere-based assays have been reported for respiratory viruses ^12,13^, viruses that cause childhood exanthems ^14^, and arthropod-borne viruses ^15,16^ There are several advantages with MBA. First, since a large number of microspheres can be coated in a single reaction, the coating would be expected to be more uniformed than those of EIA, in which each well are coated separately. Second, the signal from MBA is detected inside a flow cytometer, which avoids potential external sources that may affect the measurement of the signal. For example, scratches on microtiter plates can affect the value for EIA. Third, MBA can be easily modified into a multiplex and high-throughput platform for simultaenous detection of different antigens in multiple specimens ^13,17^ Finally, there are fewer steps and reagents involved for MBA than EIA (Supplementary Figure S1). In this study, we developed and evaluated an in-house MBA for the detection of IgG against SARS-CoV-2 nucleoprotein (NP) and spike protein receptor binding domain (RBD).

## METHODS

### Serum specimens

To set the cut-off for the EIA and MBA, we retrieved 294 archived anonymous serum specimens from the clinical biochemistry laboratory collected between April and June 2018, which were used in our previous study ^18^. For assessment of specificity, we retrieved 93 sera collected before 2020, including 53 sera collected from potential organ donors between 2016 and 2018 in a study on hepatitis E in HKSAR, and from 40 influenza patients between January and September 2019.

For COVID-19 patients, serum specimens were collected 39 recovered COVID-19 patients during the convalescent phase at the infectious disease out-patient follow-up clinic at Queen Mary Hospital. To assess the time-to-seropositivity, we retrieved 161 serial serum specimens obtained from 33 of these 39 patients during hospitalization. This study has been approved by the HKU/HA HKW Institutional Review Board (UW 13-265 and UW 18-141). Written informed consent was obtained from all COVID-19 patients.

### Cloning, purification, and biotinylation of recombinant NP and spike protein RBD of SARS-CoV-2

Cloning and purification of SARS-CoV-2 NP and spike RBD were performed as we described previously ^7^ The purified NP and spike protein RBD were biotinylated with EZ-link^TM^ Sulfo-NHS-Biotin (ThermoFisher Scientific, MA, USA). Please refer to Supplementary Methods for details.

### Enzyme immunoassay for NP and spike RBD

EIA for NP and RBD was performed as we described previously ^7^ (See supplementary Methods for details).

### Microsphere-based antibody assay

SuperAvidin™ coated microspheres (Bangs Laboratories, Indiana, USA) were washed with 500 μl of phosphate buffered saline (PBS) and 1% bovine serum albumin (BSA), and were centrifuged at 14,000 rpm at 4°C 15 minutes. Then, the microspheres were sonicated using sonicator for 30 seconds. The microspheres were coated with biotinylated NP or spike RBD, and were incubated overnight at 4°C with shaking. The microspheres were distributed onto V-bottom 96 well plates and centrifugated at 1,500 rpm and 4°C for 5 mins to remove the uncoated protein. The microspheres were then blocked with 30 μl of fetal bovine serum (FBS) for 1 h at room temperature with shaking. After blocking, FBS were removed by centrifugation at 1,500 rpm and 4°C for 5 mins, then 30 μl of serum or diluent (1% BSA, PBS) was added to each well, incubate at room temperature for 2 h with shaking. After washing, 30 μl of Alexa Fluor ® 647 AffinPure Fab fragment goat anti-human IgG, Fcγ fragment specific (5 μg/ml IgG-AF647) (Jackson ImmunoResearch, Pennsylvania, USA) was added to each well, was incubated at room temperature in the dark for 1 h with shaking. After washing, the microspheres were resuspended with 200 μl PBS and 1% BSA. Flow cytometry analysis was performed using BD LSR Fortessa analyzer (BD Biosciences, San Jose, CA, USA), and the flow cytometry data was analyzed using FlowJo v10.6.2 (FlowJo LLC, Ashland, OR, USA) (Supplementary Figure S2).

### Statistical analysis

Statistical analysis was performed using PRISM 6.0. We compared categorical variables using Fisher’s exact test and continuous variables using Mann-Whitney *U* test. The S/CO ratio was compared between MBA and EIA by Wilcoxon matched-pairs signed rank test. A *P* value of less than 0.05 was judged statistically significant.

## RESULTS

### Establishing the MBA

First we detemined the optimal microsphere-protein ratio for MBA. For NP, increasing the microsphere-protein ratio from 1:1 to 1:4 resulted in higher MFI values (Figure 1A).

However, since a microsphere-protein ratio of 1:2 could already result in a high MFI value, this ratio was selected (Figure 1B). For RBD, there was no significant difference when increasing the microsphere-protein ratio from 1:1 to 1:4 (Figure 1C). Therefore, we have selected a microsphere-protein ratio of 1:1 for RBD (Figure 1D).

**Figure 1.**
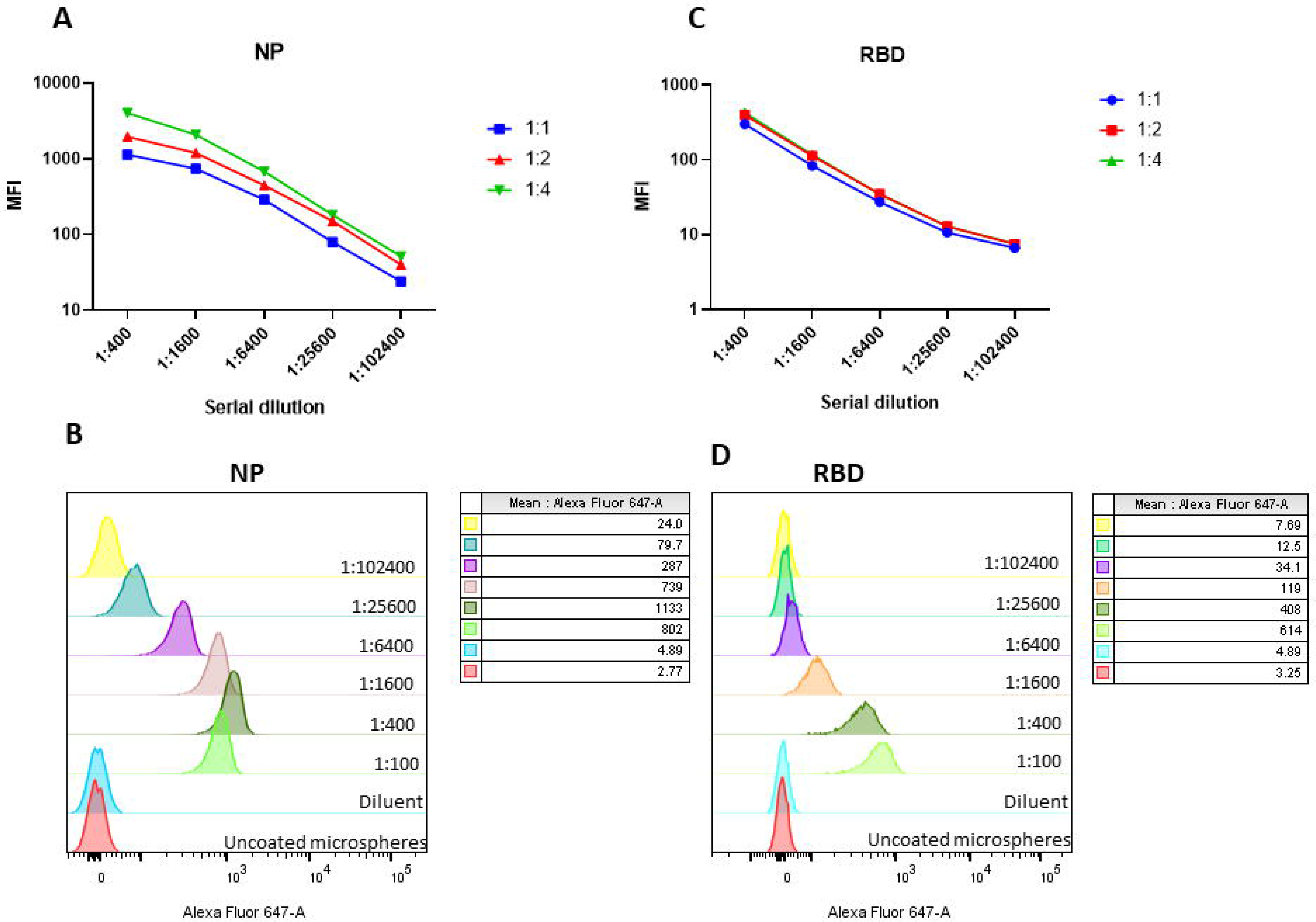
Determination of optimal microsphere-antigen ratio for microsphere-based assay. Serum from a COVID-19 patient was used. The mean fluorescent intensity at different microsphere-antigen ratio are shown for A) NP, and B) RBD), and the corresponding stacked histogram of selected microsphere-antigen ratio are shown in C) NP (1:2) and D) RBD (1:1).

Next, we tested MBA at a ratio of 1:2 for NP and 1:1 for RBD with different negative controls and a positive control (Supplementary Figure S3). The negative controls include microspheres without biotinylated NP or RBD (green), diluent only control (orange), and a serum specimen collected in 2018 (blue). The positive control was a serum specimen from a COVID-19 patient (red). All 3 negative controls had an MFI of <35, while the serum specimen from the COVID-19 patient had an MFI of 1695 for anti-SARS-CoV-2 RBD IgG and 843 for anti-SARS-CoV-2 NP IgG.

Next, we determined the optimal serum dilution for MBA and EIA using serially-diluted serum specimens from a COVID-19 patient. With both NP and RBD, the MFI plateaued at 1:400 for MBA and 1:100 provided the highest OD values with EIA (Figure 2). Hence, we have used 1:400 dilution for MBA and 1:100 for EIA for subsequent evaluation.

**Figure 2.**
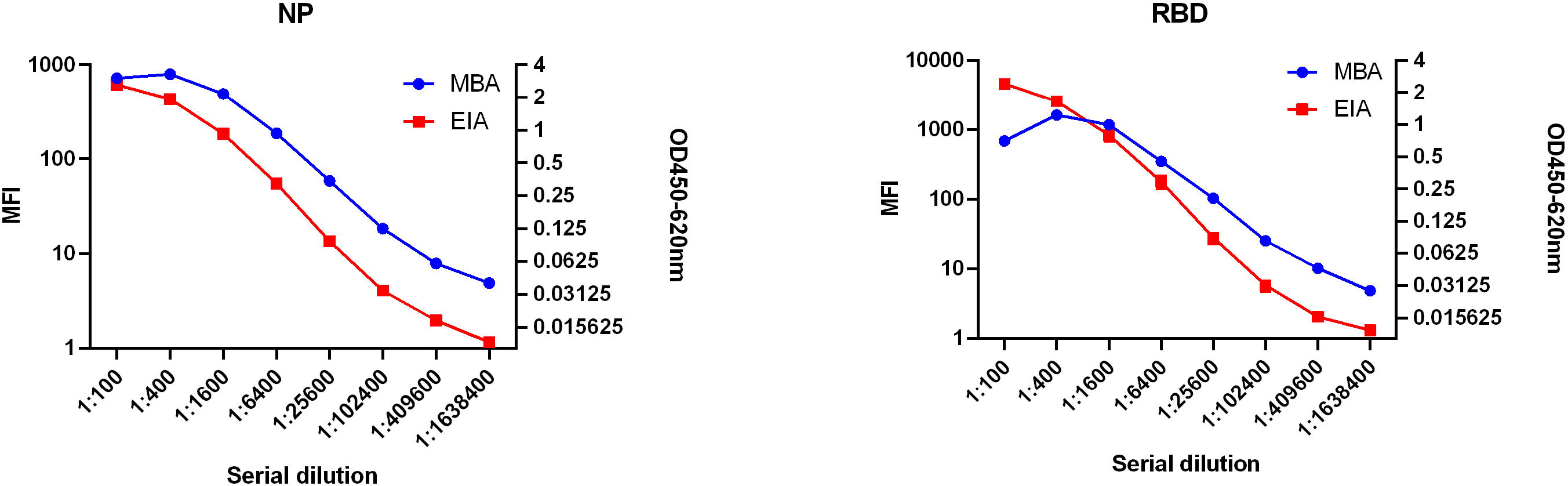
Dynamic range of MBA and EIA for A) NP, and B) RBD. Serial dilution of COVID-19 serum were used to detect anti-NP IgG and anti-RBD IgG with MBA and EIA

To determine the seropositive cutoff value for MBA, microsphere-based NP and RBD IgG assay was performed on 294 anonymous archived serum specimens collected in 2018. These anonymous serum specimens encompass all age groups from the pediatric population to those aged over 80 years (Supplementary Table S1). We first excluded outliers with >3SDs above the mean of the 294 archived anonymous serum specimens described previously ^16^. After excluding the outliers (2 for MBA anti-NP, 1 for MBA anti-RBD, 4 for EIA anti-NP, and 4 for EIA anti-RBD), the seropositive cutoff MFI (for MBA) or OD (for EIA) was then set as 3 SD above the mean of the remaining specimens. The seropositive cutoff values of MBA was 111.8 for anti-NP-IgG and 51.2 for anti-RBD IgG. The cuoff values of EIA was 0.58 for anti-NP-IgG and 0.54 for anti-RBD IgG.

To determine the specificity of the MBA, we retrieved 93 archived serum from organ donors collected between 2016 and 2018 (n=53), and from patients with influenza virus infection between January and September 2019 (n=40) (Figure 3). The specificity was 100% (93/93, 95% CI 96-100%) for anti-NP IgG and 98.9% (92/93; 95% CI 94.2-100%) for anti-RBD IgG of all negative controls (organ donors and influenza patients). Subgroup analysis showed that for organ donors, the specificity was 100% (53/53, 95% CI: 93.3-100%) for anti-NP and 98.1% (52/53; 95% CI, 89.9-100%) for anti-RBD IgG; for influenza patients, the specificities of MBA anti-NP and anti-RBD IgG were both 100% (40/40; 95% CI, 91.9-100%).

**Figure 3.**
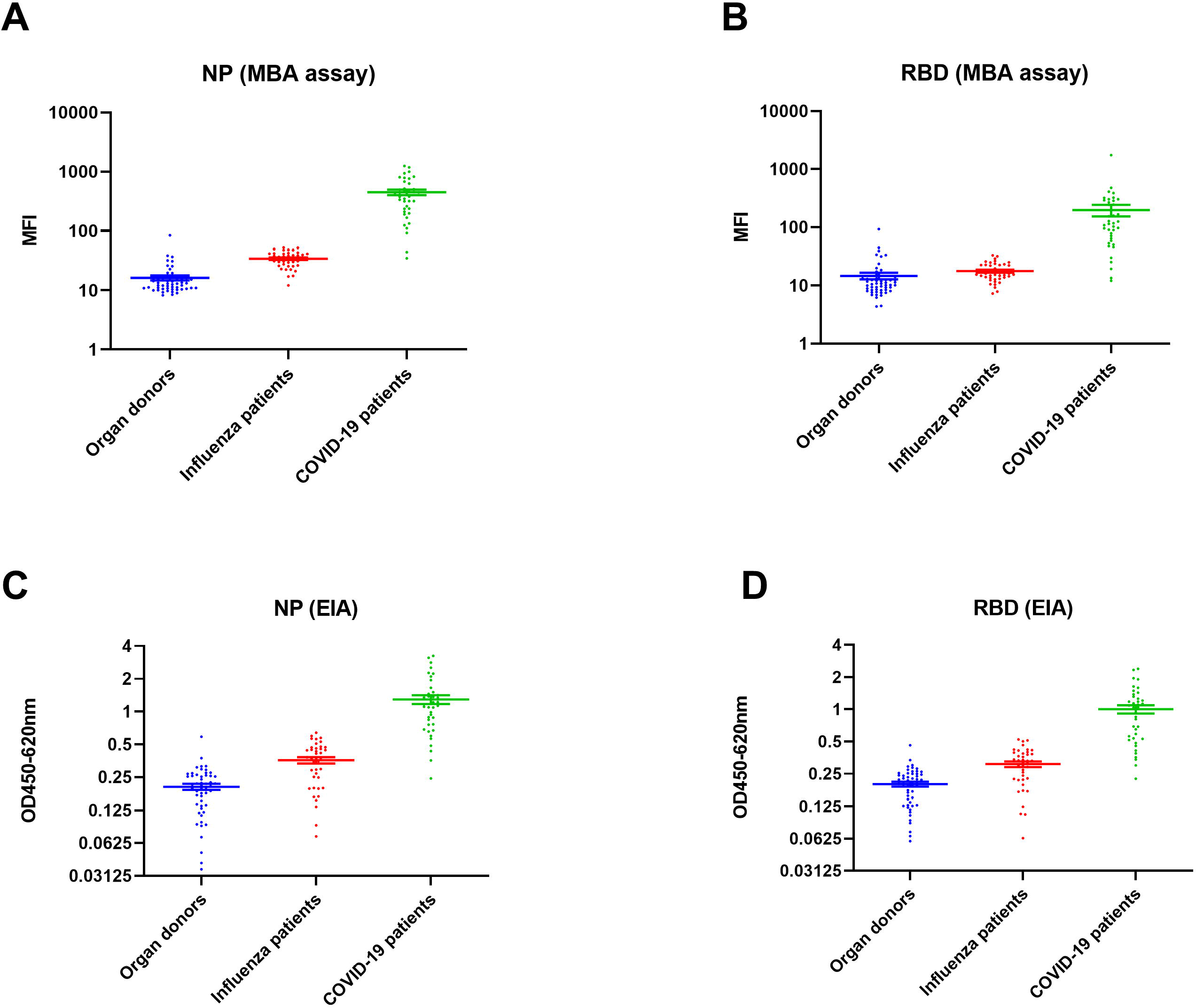
A and B) Comparison of MFI between 39 COVID-19 patients, 40 influenza patients, and 53 organ donors for A) NP and B) RBD. C and D) Comparison of OD values between COVID-19 patients, influenza patients, and organ donors for C) NP and D) RBD.

### Seropositive rate and time-to-seropositivity

Using the conditions optimized in the previous section, we determined the seropositive rate of the 39 recovered COVID-19 patient using MBA, including 16 male and 23 female patients. The median age was 57 years (range 20 to 87 years old). Of the 39 patients, 8 patients (20.5%) had severe illness requiring oxygen supplementation. The blood specimens were collected at the follow-up out-patient clinic at a median of 44 days after symptom onset (interquatile range 28 to 53 days). The seropositive rate was 89.8% (35/39) for anti-NP and 79.5% (31/39) for anti-RBD IgG.

### Time-to-seropositivity

Next, we compared the time-to-seropositivity between MBA and EIA for 33 COVID-19 patients with serial samples available during hospitalization. Out of 33 patients, 9 (27.3%) patients had anti-NP IgG detected earlier by MBA, compared to 1 (3%) detected earlier by EIA; and 9 (27.3%) patients had anti-RBD detected earlier by MBA, compared to 4 (12%) by EIA.

The time-to-seropositivity of MBA was shorter than that of EIA for both anti-NP (median time-to-sepositivity, 10 vs 12 days; hazard ratio for time-to-seropositivity, 1.41; 95% confidence interval, 0.89-2.47, P=0.1546) and anti-RBD (median time-to-sepositivity: 13 vs 14 days; hazard ratio for time-to-seropositivity: 1.05; 95% CI, 0.62-1.79, P=0.8655), though not reaching statistical significance (Figure 4).

**Figure 4.**
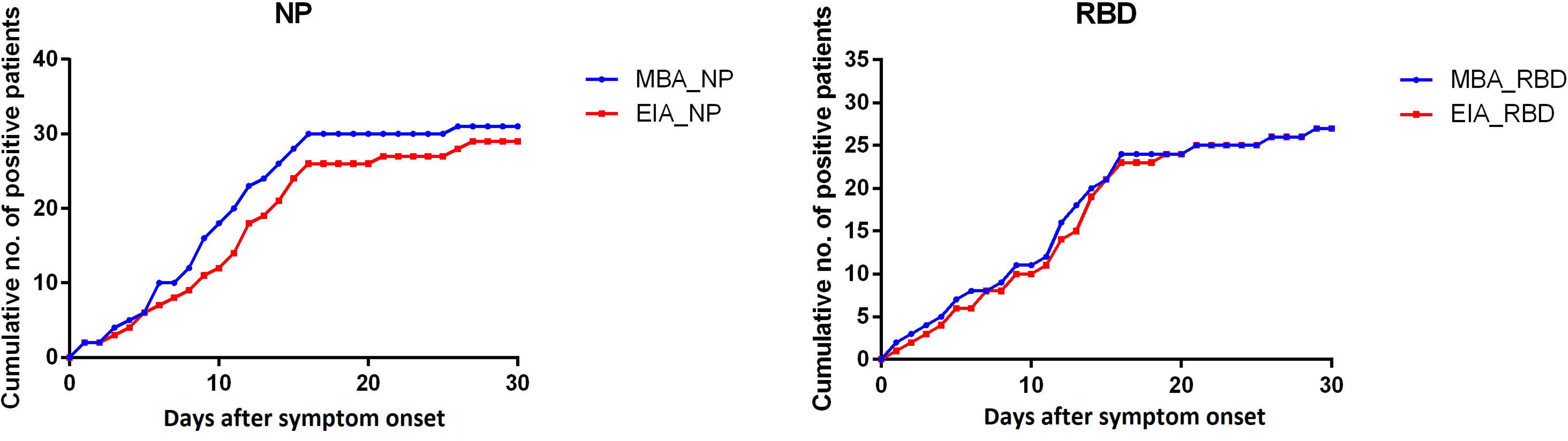
Cumulative count of seropositive COVID-19 patient specimens detected by MBA and EIA

### Comparison of S/CO

Next, we compared the signal-to-cut-off (S/CO) ratio (Figure 5). Among the convalescent serum specimens of the 39 COVID-19 patients, the S/CO ratio was significantly higher for MBA than that of EIA for both NP (median S/CO, 3.39 vs 1.95; P<0.0001) and RBD (median S/CO, 2.23 vs 1.92; P=0.0001). Among the 93 negative controls (organ donor and influenza patients), the S/CO ratio was significantly lower for MBA than that of EIA for both NP (median S/CO, 0.18 vs 0.43; P<0.0001) and RBD (median S/CO, 0.25 vs 0.43; P<0.0001). There were fewer specimens that have signal values within the equivocal range (S/CO 0.9-1.1) for MBA than those in EIA (Table 1). In particular, there were significantly fewer within the equivocal range for MBA in the negative control group for anti-NP IgG (0% [0/93] vs 8.6% [8/93]; P=0.0067).

**Figure 5.**
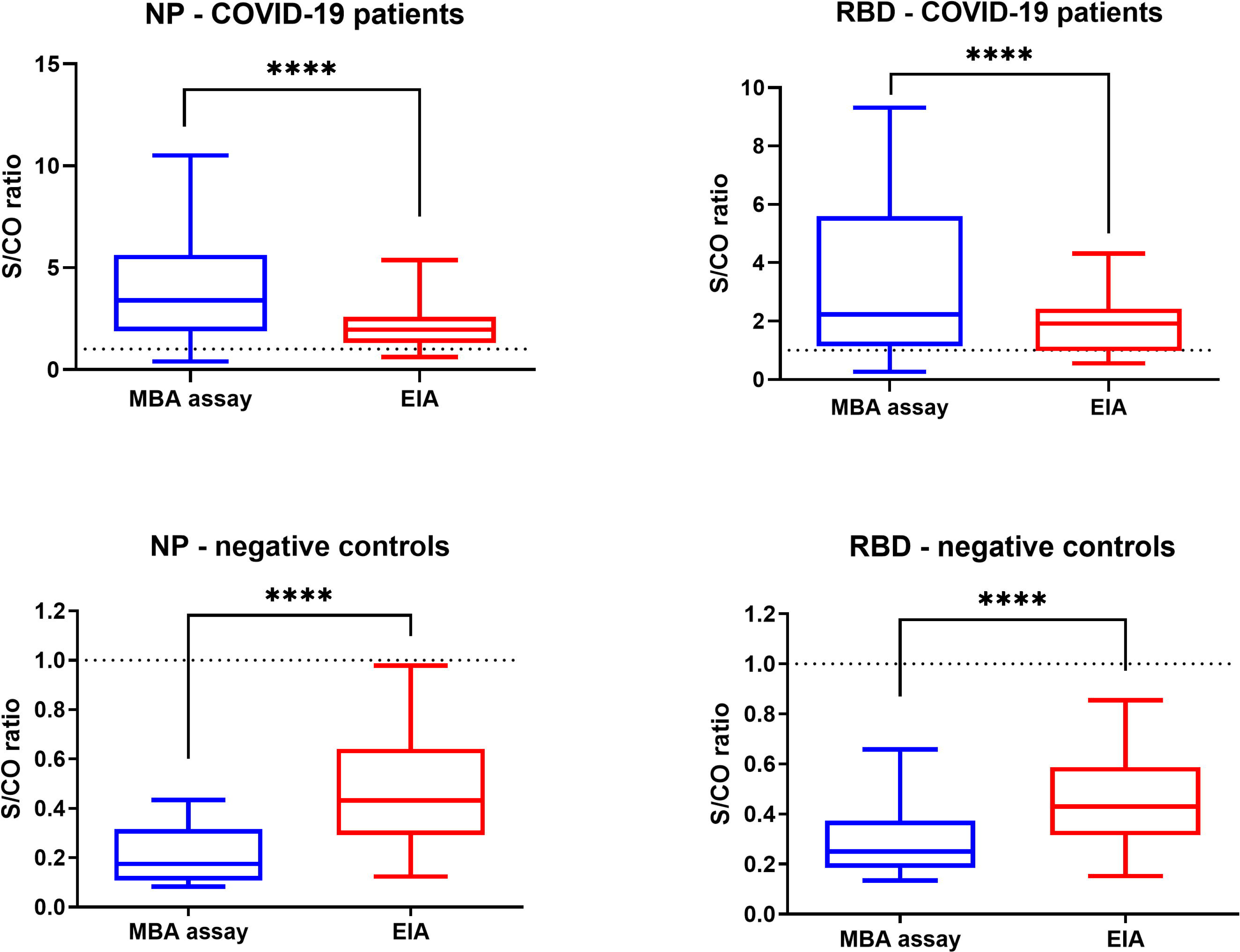
Comparison of signal-to-cut-off (S/CO) ratio between MBA and EIA for the serum specimens collected during the convalescent phase of 39 patients. ****, P≤0.0001

**Table 1.** Comparison of S/CO between MBA and EIA.

|  | Specimens collected at outpatient clinic<br>(n=39) |  | Negative controls<br>(organ donors + influenza patients)<br>(n-93) |  |
| --- | --- | --- | --- | --- |
| **MBA** | NP | RBD | NP | RBD |
| S/CO >1.1 | 34 (87.2%) | 29 (74.4%) | 0 | 1 (1.1%) |
| S/CO 0-9-1.1 | 2 (5.1%) | 5 (12.8%) | 0 | 1 (1.1%) |
| S/CO <0.9 | 3 (7.7%) | 5 (12.8%) | 93 (100%) | 91(97.8%) |
| **EIA** | NP | RBD | NP | RBD |
| S/CO >1.1 | 32 (82.1%) | 26 (66.7%) | 0 | 0 |
| S/CO 0-9-1.1 | 3 (7.7%) | 6 (15.4%) | 8 (8.6%) | 5 (12.8%) |
| S/CO <0.9 | 4 (10.3%) | 7 (18%) | 85 (91.4%) | 88 (94.6%) |

## DISCUSSION

### Principle outcomes

This study evaluated our newly developed MBA in the detection of IgG against SARS-CoV-2 NP and spike protein RBD. MBA was found to be highly specific, and had a high seropositive rate for patients with COVID-19. The time of seropositivity was shorter for MBA than that of EIA. Furthermore, when compared with EIA, MBA had a significantly higher S/CO for COVID-19 patients and a significantly lower S/CO for negative controls, and with fewer specimens within the equivocal range (S/CO 0.9-1.1). Hence, MBA is superior to EIA in the detection of IgG against SARS-CoV-2 antibody.

### Comparison with other studies

In this study, we established the seropositive cut-off values using a pre-pandemic serum panel encompassing 294 individuals from all age groups, including young children <10 years old and elderly >80 years old. This is unlike other evaluations in which healthy young adult blood donors are used as negative controls ^9^. In real life, patients often have comorbidities and many are elderlies ^19^. For some studies, the specificity was evaluted using recombinant antigen instead of serum from non-COVID-19 patients ^20^.

Many rapid lateral flow immunochromatographic assays are now commercially available. Although these immunochromatographic assays are convenient for testing, the results of these assays are poor^21,22^ or with variable sensitivity and specificity. Our flow-cytometry based MBA assay has several advantages over immunochromatographic assays. First, MBA can provide a quantitative result, while immunochromatographic assay can only give qualitative results. Therefore, MBA can assess the rise of antibody levels in a quantitative manner. Second, the interpretation of immunochromatographic assays can be difficult if the band is weakly positive. Hence there may be inter-operator difference in interpreting the results. In contrast, MBA can provide an objective readout, eliminating inter-operator difference in result interpretations.

Virus neutralization assay can detect antibodies that prevent virus from infecting cells. Since neutralization assays require the use of live SARS-CoV-2 virus, they can only be performed in biosafety level 3 laboratory. Hence, neutralization assays cannot be performed in most clinical laboratories. In contrast, the detection of IgG with recombinant virus antigens can be performed safely in biosafety level 2 clinical microbiology laboratories. Previous studies have shown that IgG correlates well with neutralizing antibody titer ^9,24^

A recent study measures anti-SARS-CoV-2 IgG level using a magnetic bead based assay ^20^. However, since a magnetic chemiluminescence analyzer is required, this may not be feasible in most clinical laboratories. In this study, our in-house MBA only requires a simple flow cytometer that is available in most clinical laboratories. Our technique can be easily applied to any laboratory with a standard flow cytometer. After simple gating and optimization, no further adjustments are required. Our method can be easily extended to other protein antigens.

### Limitations of this study

First, we only recruited adult patients. Further evaluation should be performed in pediatric patients. Second, as for all serology assays, cross-reactivity may affect the results ^9^. Even for the pre-pandemic serum, some samples can be seropositive for SARS-CoV-2 because of cross reaction with other human coronaviruses, especially from lineage B betacoronavirus. Third, samples were not tested for virus neutralization and therefore neutralizing activities of the detected IgG antibodies are unknown.

## Conclusions and implications for clinical practice and research studies

In this study, we have demonstrated that our novel flow-cytometry based MBA allowed earlier detection of anti-SARS-CoV-2 antibody among COVID-19 patients than EIA. MBA also had fewer equivocal results than EIA. A rapid and accurate diagnosis of the SARS-CoV-2 is crucial for clinicians to provide appropriate treatment to patients, to limit further spread of the virus and ultimately to eliminate another peak of pandemic risk to the public. Furthermore, our assay can be used to investigate the immune response in COVID-19 patients, establishing retrospective diagnosis especially for patients with immune-mediated diseases, determining seroprevalence in epidemiological studies, and assessing the efficacy of novel vaccines.

## Data Availability

The data will be submitted with the manuscript in different files

## ACKNOWLEDGEMENT

This study was partly supported by the donations of Richard Yu and Carol Yu, May Tam Mak Mei Yin, the Shaw Foundation Hong Kong, Michael Seak-Kan Tong, Respiratory Viral Research Foundation Limited, Hui Ming, Hui Hoy and Chow Sin Lan Charity Fund Limited, Chan Yin Chuen Memorial Charitable Foundation, Marina Man-Wai Lee, the Hong Kong Hainan Commercial Association South China Microbiology Research Fund, the Jessie & George Ho Charitable Foundation, Perfect Shape Medical Limited, and Kai Chong Tong; and funding from the Health and Medical Research Fund (grant no. COVID190124), the Food and Health Bureau, The Government of the Hong Kong Special Administrative Region; the National Program on Key Research Project of China (grant no. 2020YFA0707500 and 2020YFA0707504); the Consultancy Service for Enhancing Laboratory Surveillance of Emerging Infectious Diseases and Research Capability on Antimicrobial Resistance for Department of Health of the Hong Kong Special Administrative Region Government. The funding sources had no role in the study design, data collection, analysis, interpretation, or writing of the report.

## AUTHOR’S CONTRIBUTIONS

KKWT, CHYF designed the study, KKYT, CHYF and LLC acquired the data. KKWT, CHYF carried out the statistical analysis. All authors interpreted the data, revised the manuscript critically for important intellectual content and approved the final report.

## SUPPLEMENTARY INFORMATION

### SUPPLEMENTARY METHODS

#### Cloning and purification of (His)6-tagged recombinant receptor binding domain (RBD) and nucleocapsid protein (NP) of SARS-CoV-2

Cloning and purification of SARS-CoV-2 NP and spike RBD were performed as we described previously [7]. Briefly, the genes encoding the spike RBD (amino acid residues 306 to 543 of the spike protein) and full length NP of SARS-CoV-2 were codon-optimized, synthesized and cloned into the *NdeI* site and *XhoI* site of expression vector pET-28b(+) (Novagen, Madison, WI, USA) in frame respective and upstream of the series of six histidine residues. The recombinant RBD and NP were expressed and purified using the Ni2^+^-loaded HiTrap Chelating System (GE Healthcare, Buckinghamshire, UK) according to the manufacturer’s instructions. The purity of RBD and NP was assessed by sodium dodecyl sulfate-polyacrylamide gels (SDS-PAGE) and western blotting.

#### Biotinylation of proteins

Purified NP and spike protein RBD was diluted in PBS to 2 mg/ml and was dispensed into glass tubes on ice. 2.2 mg of EZ-link^TM^ Sulfo-NHS-Biotin (ThermoFisher Scientific, MA, USA) was dissolved in 0.5 ml sterile H2O after equilibrating to room temperature. Then, 30 μL of biotin was added to 1 ml of diluted recombinant protein and was kept on ice for 2 hours. Biotinylated recombinant proteins were dialysed using Slide-A-Lyzer^TM^ Dialysis Cassettes (20 Kd for NP and 10 Kd RBD) (ThermoFisher Scientific, MA, USA) to remove unbound biotin. The dialyzed proteins were centrifuged at 12,000 rpm for 10 min under 4 to eliminate denatured protein. The protein was stored with 50% glycerol 23 under −80. The quantity of biotinylated proteins was determined using Bradford assay.

#### Enzyme immunoassay for NP and spike RBD

EIA for NP and RBD was performed as we described previously [7]. Briefly, 96-well immunoplates (Nunc Immuno modules; Nunc, Denmark) were coated with 100 μl/well (0.1 μg/well) of SARS-CoV-2 NP or spike RBD in 0.05 M NaHCO3 (pH 9.6) overnight at 4°C and then followed by incubation with a blocking reagent. After blocking, 100 ⎧L heat-inactivated serum samples at 1:100 dilution was added to the wells and incubated at room temperature for 1 h. The attached antibodies were detected using horseradish-peroxidase (HRP)-conjugated goat anti-human IgG antibody (Invitrogen, Thermo Fisher Scientific, Waltham, MA, USA). The reaction was developed by adding diluted 3,3’,5,5’-tetramethylbenzidine (TMB) single solution and stopped with 0.3 N sulfuric acid (H2SO4). The optical density (OD) was read at 450-620 nm. A single positive sample was included in each run as positive control. An archived anonymous sample from 2018 used in our previous study was used as negative control [11]. To determine the cutoff value for positivity, the mean value of 290 anonymous archived serum specimens from 2018 plus 3 standard deviations was used as the cutoff. The cutoff values are: anti-NP IgG, 0.58; anti-NP anti-RBD IgG 0.54.

### SUPPLEMENTARY TABLES

**Supplementary Table S1.** Number of anonymous archived serum specimens in each age group

| **Age group (years)** | **2018**<br>**Apr-Jul** |
| --- | --- |
| 0-9 | 32 |
| 10-19 | 32 |
| 20-29 | 32 |
| 30-39 | 32 |
| 40-49 | 32 |
| 50-59 | 32 |
| 60-69 | 32 |
| 70-79 | 32 |
| 80 or above | 38 |
| Total | 294 |

### SUPPLEMENTARY FIGURE

**Supplementary Figure S1.**
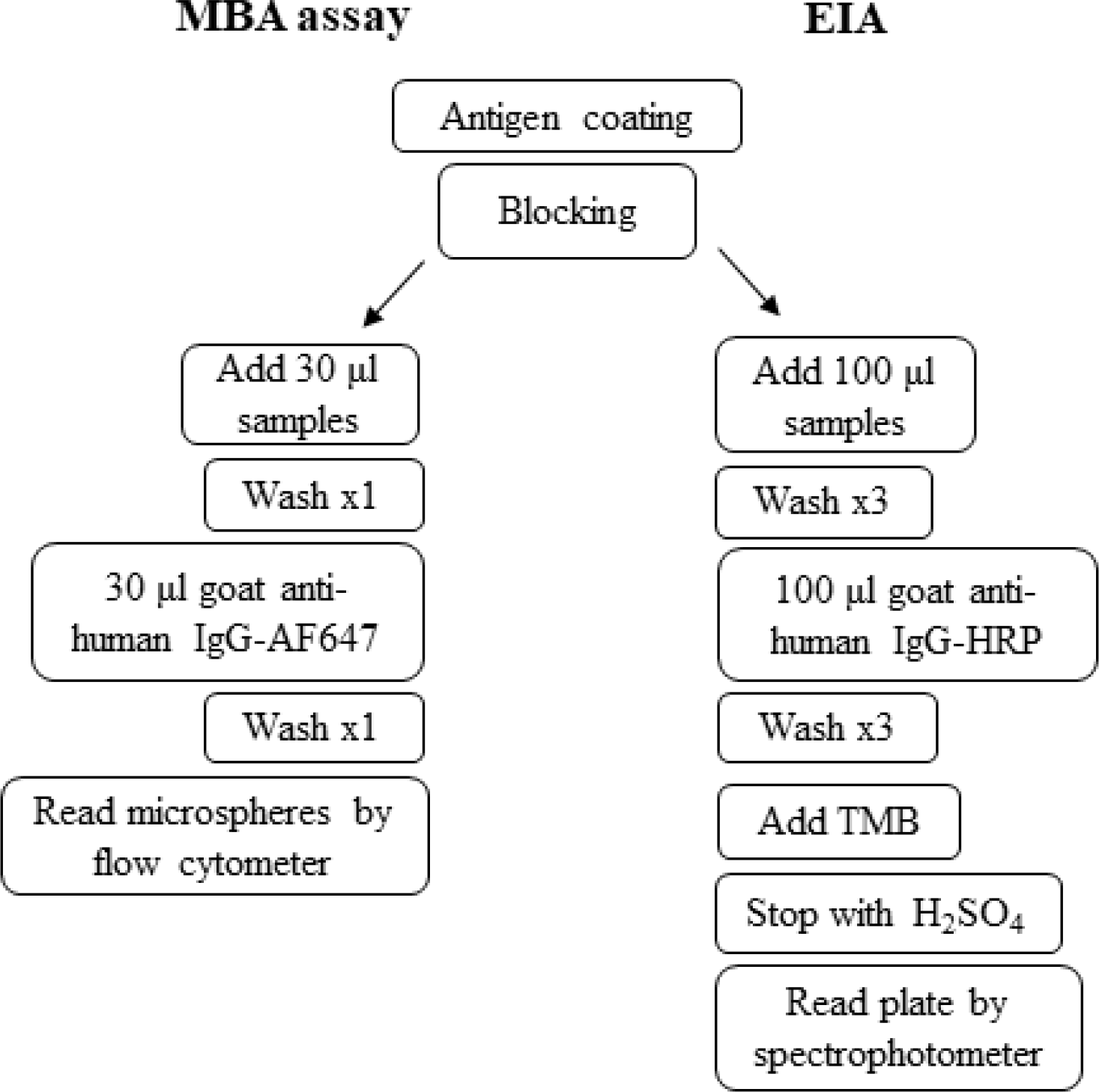
Flow chart of different steps in MBA assay and EIA

**Supplementary Figure S2.**
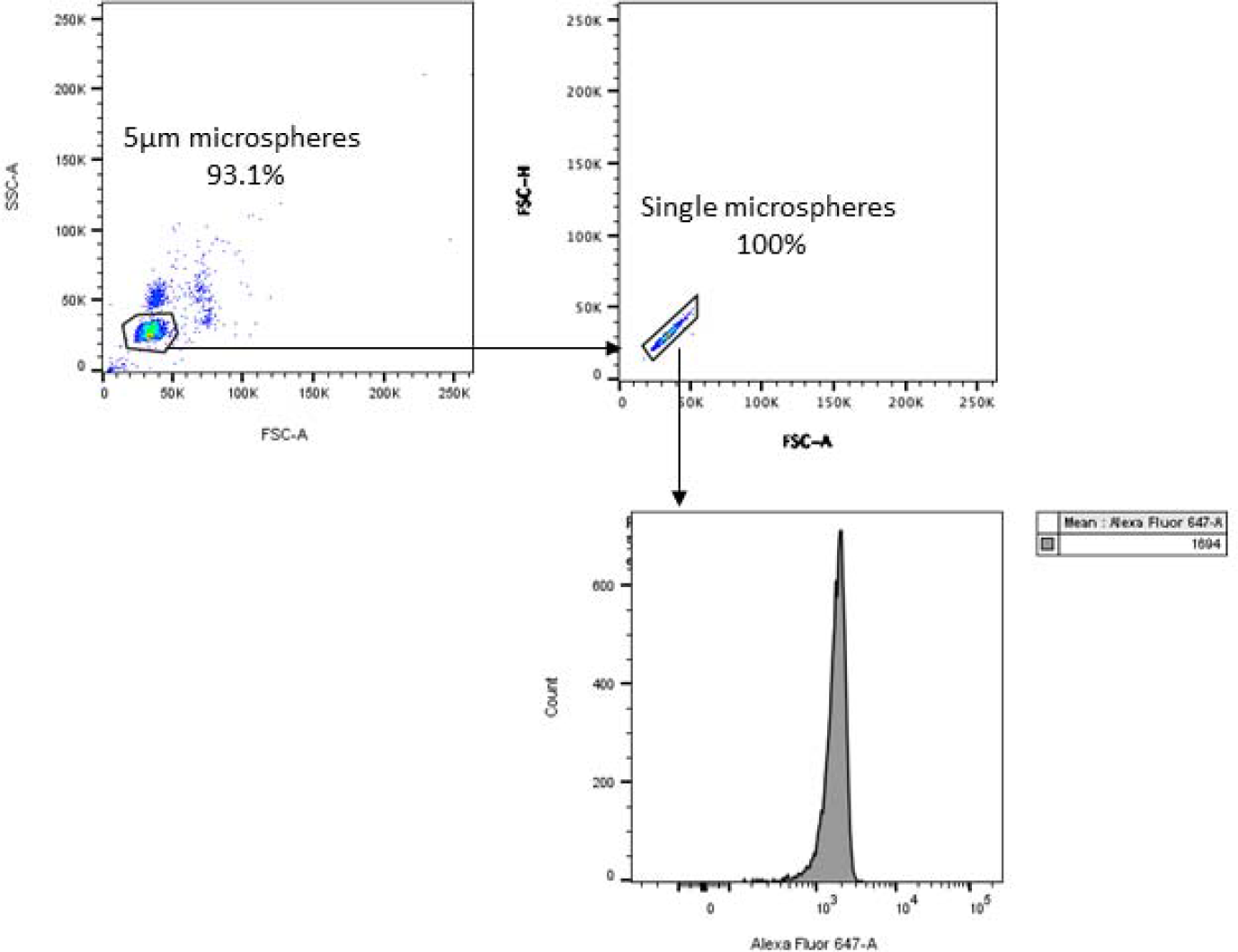
Representative flow cytometry images of the analysis of anti-NP or anti-RBD IgG.

**Supplementary Figure S3.**
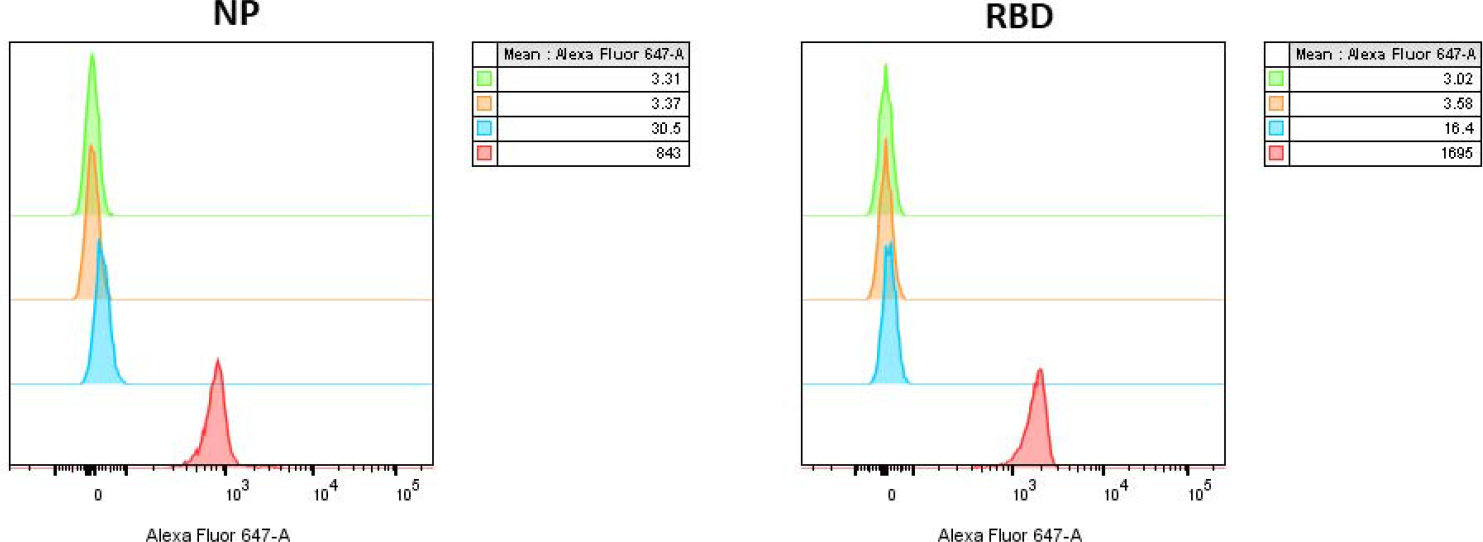
Validation of MBA assay with negative and positive controls. Streptavidin microspheres were coated with biotinylated RBD overnight at 4°C and blocked with FCS. After 1 h blocking, microspheres were washed with 1% BSA. Serum or diluent (1% BSA) were added to the microspheres and incubated for 2 hours at room temperature. Then, microspheres were washed once and goat anti-human IgG-AF647 were added. After 1 h incubation at room temperature, microspheres were washed with PBS, 1% BSA and microsphere are ready for flow cytometric analysis. The negative controls include microsphere without protein coating (green), no serum (diluent only) (orange), and a serum specimen collected in 2018 (blue). The positive control was a serum specimen from a patient with COVID-19 (red).

